# THE CARDIO-PAL SCORE – A NOVEL PROGNOSTICATION TOOL FOR 6-MONTH MORTALITY IN HEART FAILURE INPATIENTS

**DOI:** 10.64898/2026.09.28.26364242

**Authors:** Hilary J. Bews, James W. Tam, Samuel Quan, Casandra Dolovich, Klara Whalen, Sarah-Jane Beaudry, Stephanie Butterworth, Anika Southam, Jana L. Pilkey

**Author notes:** Address for correspondence: Hilary J. Bews MD, B.Sc. FRCPC St. Boniface Hospital 409 Tache Ave. Winnipeg, Manitoba, Canada R2H 2A6.

## Abstract

**Background:** Patients with heart failure (HF) experience high mortality, yet survival estimates remain challenging. Available prognostic tools commonly predict survival beyond 1-year, limiting goals of care discussions and enrollment on palliative care programs. The objective of this project was to develop a novel prognostic score to better predict 6-month mortality.

**Methods:** Patients with HF admitted to the St. Boniface Hospital (Winnipeg, Canada) between January 1, 2019 and January 1, 2024 were identified. Patient characteristics, imaging parameters, laboratory values, and functional assessments (Palliative Performance Scale; PPSv2) were collected. Patients were randomly divided into development (80%) and validation (20%) cohorts. Penalized LASSO regression was used to select variables most predictive of 6-month mortality in the development cohort. Retained variables were incorporated into a multivariable logistic regression model to derive the CARDIO-PAL score, the performance of which was assessed in the holdout validation cohort.

**Results:** Characteristics were similar between the development (n = 2036) and validation (n = 508) cohorts, with 6-month mortality rates of 29% and 31%, respectively. 8 candidate variables were retained: age (adjusted odds ratio [aOR] 1.01), PPSv2 (aOR 2.08), systolic blood pressure (aOR 1.14), discontinued use of angiotensin-converting enzyme inhibitor or angiotensin II receptor blocker (aOR 2.58), beta-blocker (aOR 1.59), and mineralocorticoid receptor antagonist (aOR 2.67), right ventricular systolic pressure (aOR 2.06), and urea (aOR 2.13). The CARDIO- PAL score demonstrated good discrimination, with an area under the curve of 0.80 (95% confidence interval 0.75 – 0.84) and good calibration (slope 0.84, intercept -0.04). 6-month mortality across low, intermediate, and high-risk groups was 2%, 15%, and 54%, respectively.

**Conclusion:** The CARDIO-PAL score is a novel tool to predict 6-month mortality in HF inpatients. A collaborative approach integrating CARDIO-PAL with other prognostic scores and serial clinical assessments has the potential to promote patient-centered care and improve referral to palliative care programs.

## Introduction

Advanced (stage D) heart failure (HF) is defined by severe symptoms at rest or with minimal exertion, recurrent hospitalizations, medication refractoriness, and consideration of advanced therapies or palliative care.^1^ It is characterized by a non-linear disease trajectory and a high short-term mortality of 80% at five years.^2,3^ Patients with HF report high physical, psychosocial, and spiritual needs.^3^ Accordingly, the integration of palliative care throughout all stages of HF is a Class 1 recommendation in recent European Society of Cardiology and American College of Cardiology/American Heart Association HF guidelines.^4,5^ In contrast to this recommendation, only a minority of patients with HF are referred to palliative care.^3^

Despite the high mortality associated with advanced HF, survival estimates for individual patients remain challenging. Although prognostic tools exist, clinical applicability is limited by invasive or obscure variables and a lack of generalizability to multimorbid patients, because many of the tools were derived from highly selective patient populations. Furthermore, relatively few scores were developed to predict short term (≤ 6-month) mortality, which can limit goals of care conversations and patient access to palliative care and hospice programs.^6^ Additionally, most available HF prognostic scores do not incorporate functional parameters, despite a strong association with mortality.^7^

The objective of this study was to develop and validate a novel, generalizable and practical prognostic score, which includes a functional assessment, to help better define 6-month mortality in HF inpatients. Although a clear understanding of prognosis is not the only barrier to goals of care discussions, an accurate prognostic tool to predict short term mortality has the potential to promote patient-centered care, and help support decision making around hospice and palliative care referrals.

## Methods

### Patient Population

This retrospective cohort study included all patients ≥ 18 years admitted to the St. Boniface Hospital (tertiary academic hospital in Winnipeg, Manitoba, Canada) with a primary diagnosis of HF between January 1, 2019 and January 1, 2024. Sample size was determined by the number of records available. Data collection occurred between April 7, 2025 and September 27, 2025. For patients with multiple hospitalizations, the most recent visit within the study period was used. Patients were excluded for the following reasons: non-Manitoba medical record number, left ventricular assist device (LVAD), additional privacy features that limited chart review, lack of clinical heart failure upon chart review, inability to access electronic patient chart, recent heart transplant, or provision of medical assistance in dying (MAID). All comorbidities and range of ejection fractions were permitted as a reflection of a real-world population. Ethics approval was obtained from the University of Manitoba, Health Research Ethics Board, Bannatyne Campus (HS26709[H2024:338]) and the St. Boniface Hospital Research Review Committee (RRC/2024/2191).

### Study Variables

Patient demographics collected included age, sex, and body mass index (BMI). BMI was calculated using the weight closest to hospital discharge. Clinical variables collected included triage vital signs, major comorbidities, smoking status, number of unplanned hospital visits in the preceding 12 months, and past or current use of cardiac medications. Triage vital signs recorded included heart rate (HR), systolic blood pressure (SBP), respiratory rate (RR), oxygen saturation, and use of oxygen. Comorbidity variables collected were the presence of hypertension, diabetes, chronic kidney disease, liver disease, cirrhosis, cancer, chronic obstructive pulmonary disease (COPD), dementia, peripheral arterial disease, cerebrovascular disease (history of transient ischemic attack, stroke, endarterectomy, or carotid or vertebral artery occlusion), significant coronary artery disease (on angiogram, prior myocardial infarction, percutaneous coronary artery intervention, or coronary artery bypass graft surgery), coronavirus disease 2019 (COVID-19), and atrial fibrillation. Cardiac medications recorded included use of angiotensin-converting enzyme inhibitor (ACEI), angiotensin II receptor blocker (ARB), angiotensin receptor-neprilysin inhibitor (ARNI), beta-blocker, mineralocorticoid receptor antagonist (MRA), and sodium-glucose cotransporter 2 inhibitor (SGLT2). Unplanned hospital visits included emergency department visits and hospital admissions.

Functional variables collected included the discharge physiotherapy score (meters ambulated), presence of wounds/ulcers, and Palliative Performance Scale version 2 (PPSv2). The PPS was originally developed by the Victoria Hospice Society for patients with cancer, but was later adapted, and validated to a palliative HF population.^8–10^ It is a functional assessment score based on ambulation, activity, self-care, intake and level of consciousness (Supplementary Figure 1). A percentage is assigned in 10% increments between 0% (death) to 100% (fully functional) based on the score in each category. PPSv2 was assigned on admission, or at the closest point to admission possible to clearly reflect a true estimate of functionality (i.e. after extubation and removal of sedation if in a critical care setting). Written permission to use the PPSv2 was obtained from Victoria Hospice.

Laboratory values were collected at admission and discharge and included: sodium (mmol/L), potassium (mmol/L), bicarbonate (mmol/L), albumin (g/L), urea (mmol/L), creatinine (µmol/L), gamma-glutamyl transferase (GGT; U/L), alanine aminotransferase (ALT; U/L), white blood count (X10^9^/L), absolute neutrophil count (X10^9^/L), absolute lymphocyte count (X10^9^/L), lymphocyte percentage, hemoglobin (g/L), platelets (X10^9^/L), and N-terminal-pro B-type natriuretic peptide (NT-proBNP; pg/mL). Of note, NT-proBNP was only available to cardiologists in our center prior to 2020, before becoming universally available.

Finally, cardiac imaging parameters were collected from available echocardiogram reports, closest to the date of discharge. Parameters obtained included left ventricular ejection fraction (LVEF), left ventricular internal diameter in diastole (LVIDd), right ventricular dysfunction (normal, less than severe, severe), right ventricular systolic pressure (RVSP, mmHg), and right atrial pressure (normal [< 5 mmHg], less than severe [5 – 15 mmHg], severe [>15 mmHg]). A second cardiologist specializing in echocardiography, reviewed source images when discrepancies in the data existed. The outcome variable of interest was short-term mortality, defined as death occurring within 6- months of hospital discharge.

## Statistical Analysis

### Splitting the data

The sample was randomly divided using a Statistical Analysis System (SAS version 9.4) and corresponding method, stratified random sampling (PROC SURVEYSELECT). The development cohort comprised 80% of the total sample, and the independent holdout validation cohort comprised the remaining 20%. Cohorts were created following comprehensive data quality checks and preprocessing. The validation cohort remained sealed until final model evaluation. Descriptive statistics were presented for both the development and validation cohorts as percentages for categorical variables and means with standard deviations (SD), medians, and interquartile ranges (IQR) for continuous variables, when appropriate.

Analyses were conducted in accordance with recommendations from the Transparent Reporting of a multivariable prediction model for Individual Prognosis Or Diagnosis (TRIPOD) Statement for reporting prediction model development and validation.^11^

### Missing Data and Variable Selection

Four candidate variables had missingness greater than 25% including laboratory variables NT-proBNP, ALT, GGT, and echo variable RVSP. Missing data were handled using single imputation based on fully conditional specification with predictive mean matching, including all candidate predictors and the outcome in the imputation model. Continuous variables with skewed distributions were log-transformed prior to imputation. During exploratory model development, a subset of the development cohort (60% of the total sample) was used for initial predictor selection, with final model estimation performed using the full development cohort (80%). Penalized regression Least Absolute Shrinkage and Selection Operator (LASSO; PROC GLMSELECT) with 10-fold cross-validation was used for variable selection. Our data included over 50 candidate variables. The LASSO method functions to reduce model overfitting and improve ease of use by shrinking regression coefficients and selecting a parsimonious set of predictors from a larger candidate variable set. Given the computational complexity and potential instability of combining repeated multiple imputation with cross-validated LASSO procedures, a single imputed dataset was used for model development. For variables in which missingness was considered potentially non-random (e.g. RVSP), a binary missing indicator was included during variable selection. Model performance was subsequently evaluated in an independent holdout validation dataset to assess predictive performance in unseen data. This process was performed separately for admission and discharge laboratory variables. Due to substantial missingness in several discharge laboratory variables (up to 75%), the final analysis was restricted to admission variables.

### Calculation of the CARDIO-PAL Score

Variables retained in the LASSO variable selection step were included in a multivariable logistic regression model to predict 6-month mortality in the development cohort. A point-based scoring system was derived from the regression coefficients to develop the Cardiac Assessment of Risk to Determine Illness Outcomes and Palliative Care (CARDIO-PAL) score. Points were assigned by dividing each regression coefficient (β) by a scaling factor of 0.25 and rounding to the nearest integer. Variables with a contribution corresponding to less than one point were assigned zero points in the final scoring system. Age did not contribute to the final score but was retained in the model as a clinically meaningful variable to preserve the influence on the coefficients of the other predictors. SBP was centered at the cohort median (130 mmHg), with 1 point assigned per 10- mmHg decrease below this value and −1 point assigned per 10-mmHg increase above this value. PPSv2 demonstrated a linear relationship with 6-month mortality and was therefore modeled as a continuous variable. Using 80% as the reference value, three points were assigned per 10% decrease in PPSv2, corresponding to a possible range of 0 to 21 points. For continuous variables, points were assigned per unit increase (e.g., 3 × log[urea]), while categorical variables contributed fixed point values based on category (e.g., −1 for current beta blocker use). The CARDIO-PAL score was calculated as the sum of points assigned to each predictor included in the final model, with higher scores corresponding to a higher risk of 6-month mortality. The corresponding formula for calculating the total CARDIO-PAL score is provided for reference.

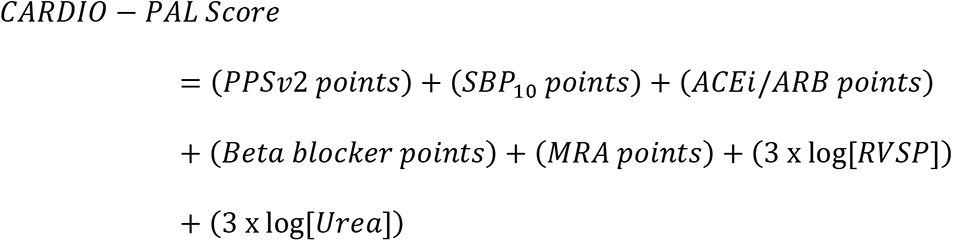

### Detailed point assignments for eacℎ predictor are provided in tℎe results section

Since RVSP is not routinely available in all clinical settings, two versions of the prediction model were developed: a full model including all predictors retained through LASSO selection, and a reduced model excluding RVSP. Both models were developed using the same modeling procedures and evaluated using identical performance metrics to assess the impact of excluding RVSP. Therefore, two versions of the CARDIO-PAL score were calculated.

For clinical interpretability, CARDIO-PAL scores were categorized into low-, intermediate-, and high-risk groups. Risk groups were defined based on predefined predicted probability ranges (<10%, 10-30%, and ≥30%)^12^ and corresponding CARDIO-PAL score ranges derived from the development cohort. Observed 6-month mortality was reported for each risk group. Kaplan-Meier survival curves were generated by CARDIO-PAL risk group to estimate cumulative all-cause mortality following discharge, with follow-up administratively censored on April 7, 2025. An expanded view of the first 180 days of follow-up was generated to illustrate early mortality patterns.

### Performance of the CARDIO-PAL

Model performance was evaluated in both the development cohort (apparent performance) and the holdout validation cohort. Discrimination was evaluated using the area under the receiver operating characteristic curve (AUC). Calibration in the validation cohort was assessed by comparing predicted and observed 6-month mortality. Internal model validation was assessed using bootstrap resampling (1,000 iterations) to estimate optimism in model performance. Optimism-corrected estimates were reported for both the AUC and calibration slope, calculated as the apparent performance minus the mean optimism. The calibration intercept (calibration-in-the-large) and slope were also evaluated via the validation cohort. Sensitivity, specificity, positive predictive value (PPV), and negative predictive value (NPV) were calculated for the original PPSv2 and the new CARDIO-PAL mortality risk score (with and without RVSP). Performance thresholds were defined a priori, using PPSv2 ≤ 40% based on previously established prognostic literature^13^ and predicted probability ≥ 0.55 based on the CARDIO-PAL score, which was selected in the development cohort to prioritize high specificity, matching that of PPSv2 ≤ 40%.

## Results

### Patient Characteristics

Among 2651 patients, 107 (4.0%) were excluded, resulting in a final sample of 2544 patients (Figure 1). The development and validation cohorts included 2036 (80%) and 508 (20%) patients, respectively.

**Figure 1:**
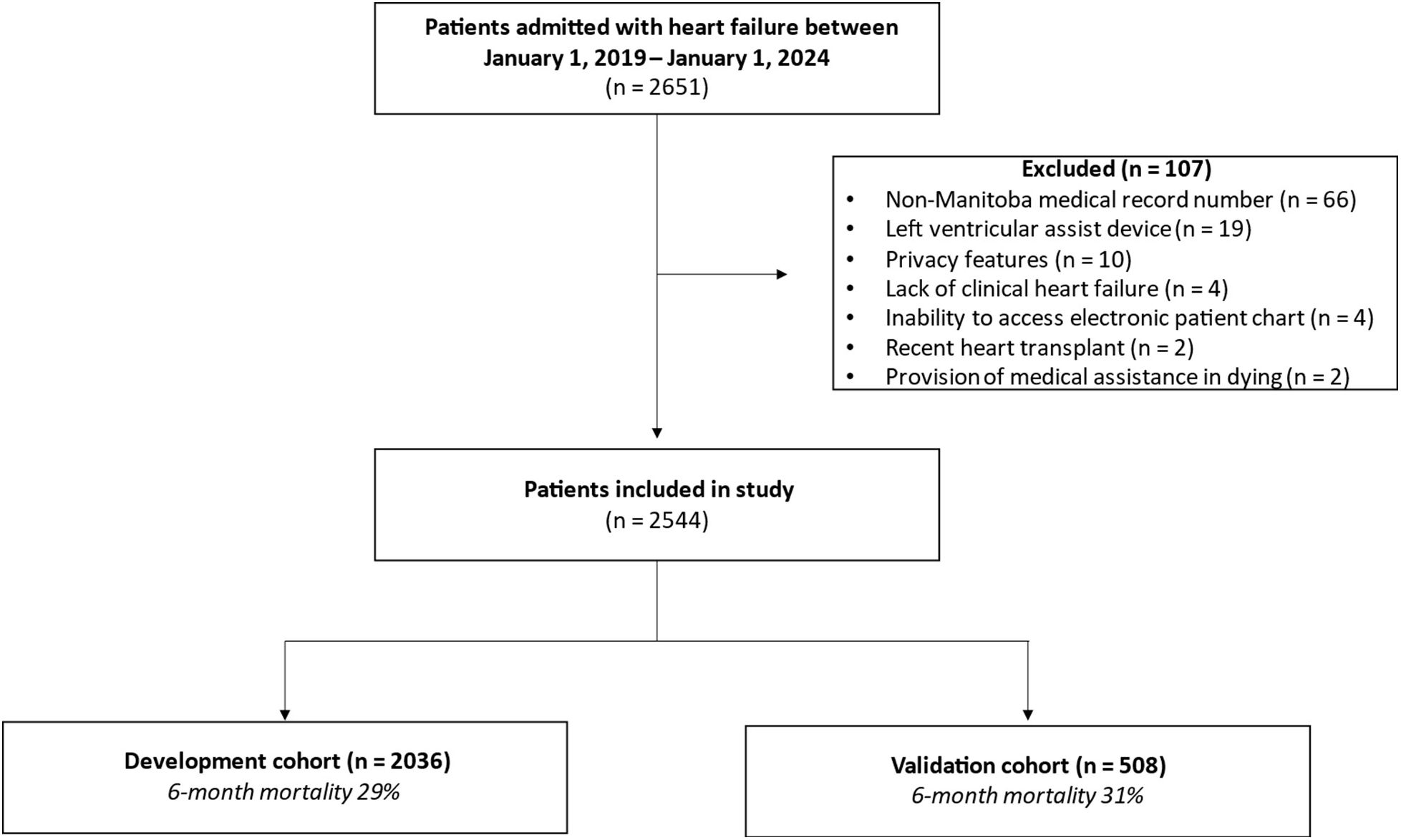
Flow diagram of patients through the study.

Baseline characteristics and candidate predictors for the development cohort are outlined in Table 1. The development cohort had a mean age of 74 years (SD 14.4 years), mean BMI of 29.4 kg/m^2^ (SD 8.6 kg/m^2^), and 43.7% were female. Common comorbidities included hypertension (74.1%), atrial fibrillation/flutter (47.1%), diabetes (46.9%), significant coronary artery disease (44.6%), chronic kidney disease (34.4%), and COPD (21.2%). Only 5% of patients were found to be COVID-19 positive at any point during their hospital stay. The median number of unplanned hospital visits in the 12 months leading up to the index hospitalization was 2 (IQR 0 – 3) and use of cardiac medications were as follows; 50.6% were currently on an ACEI/ARB, 69.8% on a beta-blocker, 26.5% on a MRA, and 13% on a SGLT2. LVEF was preserved (LVEF > 50%) in 45.8%, and reduced (LVEF ≤ 50%) in 51.1%. RV function was normal on echocardiography in 42.7%, with less than severe RV dysfunction in 39.9% and severe RV dysfunction in 13.2%. The median RVSP was 43 mmHg (IQR 35 – 53 mmHg), and right atrial pressure was < 5 mmHg in 35%, 5 - 15 mmHg in 29.2%, and > 15 mmHg in 27% of the cohort. The majority of patients were assigned a PPS ≥ 50%, with 48.7% ambulating a distance of < 150 m at discharge.

**Table 1.**
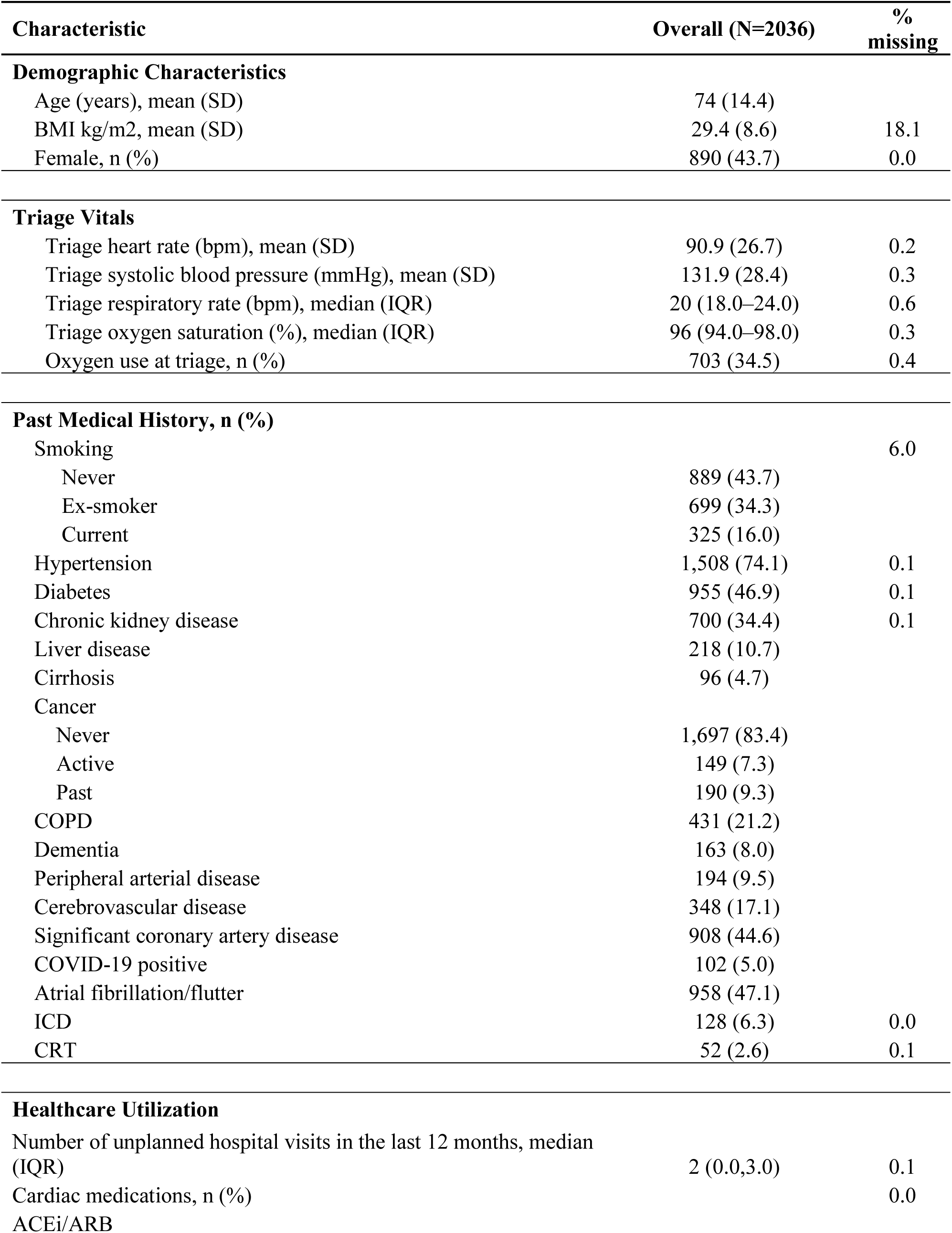
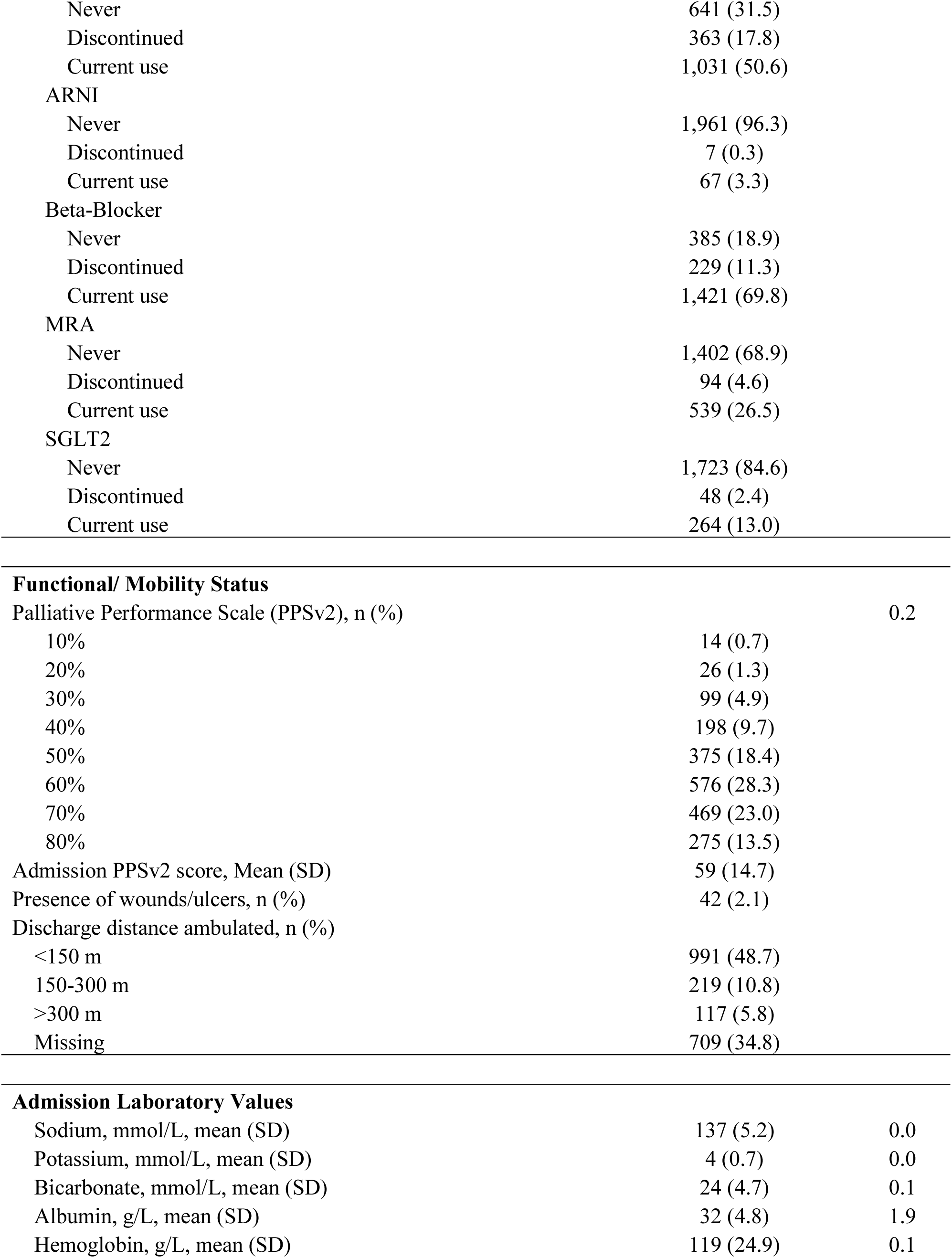
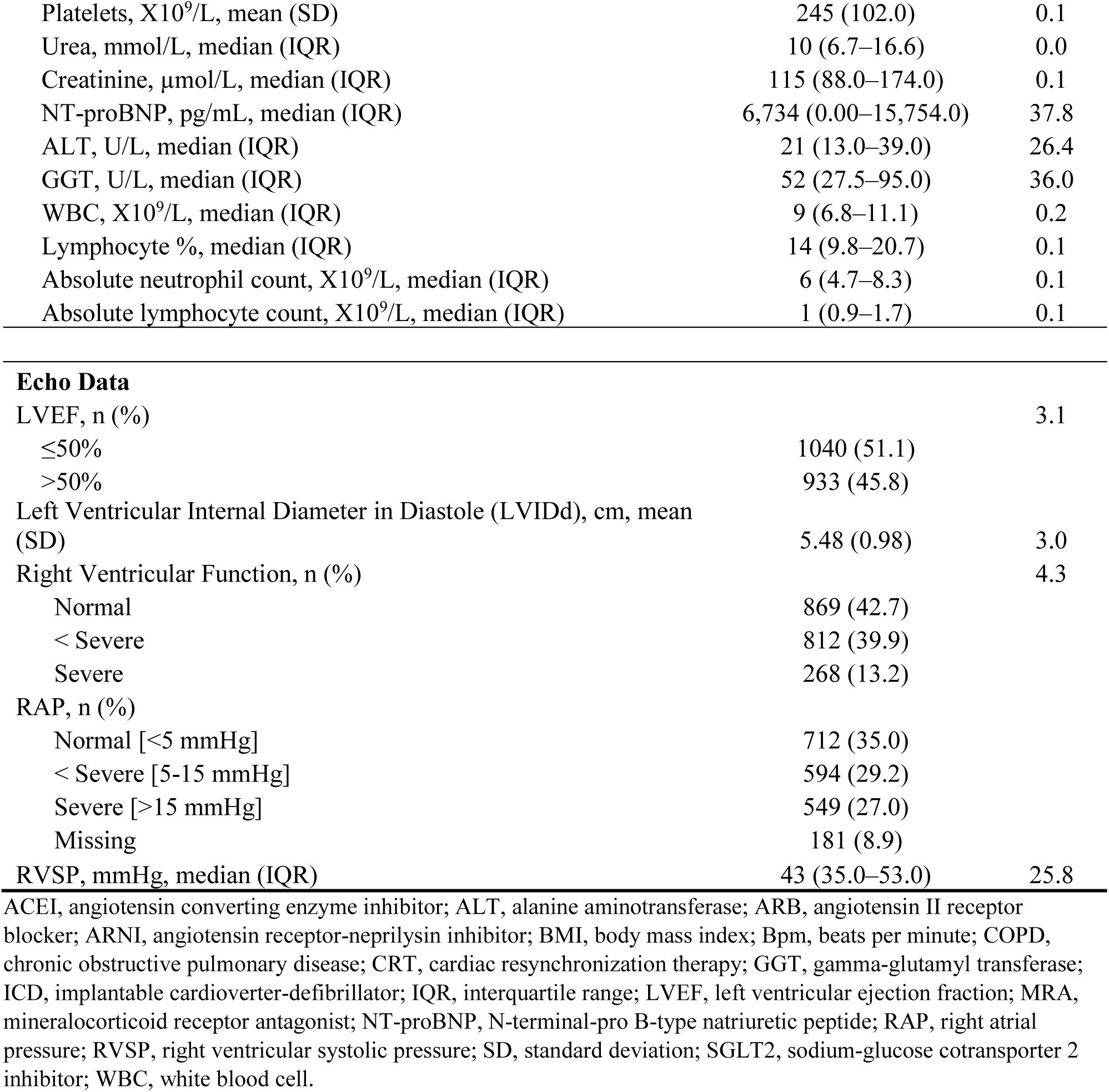
Baseline Characteristics and Candidate Predictors in the Development Cohort before Imputation.

| Characteristic | Overall (N=2036) | % missing |
| --- | --- | --- |
| **Demographic Characteristics** |  |  |
| Age (years), mean (SD) | 74 (14.4) |  |
| BMI kg/m<sup>2</sup>, mean (SD) | 29.4 (8.6) | 18.1 |
| Female, n (%) | 890 (43.7) | 0.0 |
| **Triage Vitals** |  |  |
| Triage heart rate (bpm), mean (SD) | 90.9 (26.7) | 0.2 |
| Triage systolic blood pressure (mmHg), mean (SD) | 131.9 (28.4) | 0.3 |
| Triage respiratory rate (bpm), median (IQR) | 20 (18.0–24.0) | 0.6 |
| Triage oxygen saturation (%), median (IQR) | 96 (94.0–98.0) | 0.3 |
| Oxygen use at triage, n (%) | 703 (34.5) | 0.4 |
| **Past Medical History, n (%)** |  |  |
| Smoking |  | 6.0 |
| Never | 889 (43.7) |  |
| Ex-smoker | 699 (34.3) |  |
| Current | 325 (16.0) |  |
| Hypertension | 1,508 (74.1) | 0.1 |
| Diabetes | 955 (46.9) | 0.1 |
| Chronic kidney disease | 700 (34.4) | 0.1 |
| Liver disease | 218 (10.7) |  |
| Cirrhosis | 96 (4.7) |  |
| Cancer |  |  |
| Never | 1,697 (83.4) |  |
| Active | 149 (7.3) |  |
| Past | 190 (9.3) |  |
| COPD | 431 (21.2) |  |
| Dementia | 163 (8.0) |  |
| Peripheral arterial disease | 194 (9.5) |  |
| Cerebrovascular disease | 348 (17.1) |  |
| Significant coronary artery disease | 908 (44.6) |  |
| COVID-19 positive | 102 (5.0) |  |
| Atrial fibrillation/flutter | 958 (47.1) |  |
| ICD | 128 (6.3) | 0.0 |
| CRT | 52 (2.6) | 0.1 |
| **Healthcare Utilization** |  |  |
| Number of unplanned hospital visits in the last 12 months, median (IQR) | 2 (0.0,3.0) | 0.1 |
| Cardiac medications, n (%) |  | 0.0 |
| ACEi/ARB |  |  |
| Never | 641 (31.5) |  |
| Discontinued | 363 (17.8) |  |
| Current use | 1,031 (50.6) |  |
| ARNI |  |  |
| Never | 1,961 (96.3) |  |
| Discontinued | 7 (0.3) |  |
| Current use | 67 (3.3) |  |
| Beta-Blocker |  |  |
| Never | 385 (18.9) |  |
| Discontinued | 229 (11.3) |  |
| Current use | 1,421 (69.8) |  |
| MRA |  |  |
| Never | 1,402 (68.9) |  |
| Discontinued | 94 (4.6) |  |
| Current use | 539 (26.5) |  |
| SGLT2 |  |  |
| Never | 1,723 (84.6) |  |
| Discontinued | 48 (2.4) |  |
| Current use | 264 (13.0) |  |
| Functional/ Mobility Status |  |  |
| Palliative Performance Scale (PPSv2), n (%) |  | 0.2 |
| 10% | 14 (0.7) |  |
| 20% | 26 (1.3) |  |
| 30% | 99 (4.9) |  |
| 40% | 198 (9.7) |  |
| 50% | 375 (18.4) |  |
| 60% | 576 (28.3) |  |
| 70% | 469 (23.0) |  |
| 80% | 275 (13.5) |  |
| Admission PPSv2 score, Mean (SD) | 59 (14.7) |  |
| Presence of wounds/ulcers, n (%) | 42 (2.1) |  |
| Discharge distance ambulated, n (%) |  |  |
| <150 m | 991 (48.7) |  |
| 150-300 m | 219 (10.8) |  |
| >300 m | 117 (5.8) |  |
| Missing | 709 (34.8) |  |
| Admission Laboratory Values |  |  |
| Sodium, mmol/L, mean (SD) | 137 (5.2) | 0.0 |
| Potassium, mmol/L, mean (SD) | 4 (0.7) | 0.0 |
| Bicarbonate, mmol/L, mean (SD) | 24 (4.7) | 0.1 |
| Albumin, g/L, mean (SD) | 32 (4.8) | 1.9 |
| Hemoglobin, g/L, mean (SD) | 119 (24.9) | 0.1 |
| Platelets, X10<sup>9</sup>/L, mean (SD) | 245 (102.0) | 0.1 |
| Urea, mmol/L, median (IQR) | 10 (6.7–16.6) | 0.0 |
| Creatinine, μmol/L, median (IQR) | 115 (88.0–174.0) | 0.1 |
| NT-proBNP, pg/mL, median (IQR) | 6,734 (0.00–15,754.0) | 37.8 |
| ALT, U/L, median (IQR) | 21 (13.0–39.0) | 26.4 |
| GGT, U/L, median (IQR) | 52 (27.5–95.0) | 36.0 |
| WBC, X10<sup>9</sup>/L, median (IQR) | 9 (6.8–11.1) | 0.2 |
| Lymphocyte %, median (IQR) | 14 (9.8–20.7) | 0.1 |
| Absolute neutrophil count, X10<sup>9</sup>/L, median (IQR) | 6 (4.7–8.3) | 0.1 |
| Absolute lymphocyte count, X10<sup>9</sup>/L, median (IQR) | 1 (0.9–1.7) | 0.1 |
| Echo Data |  |  |
| LVEF, n (%) |  | 3.1 |
| ≤50% | 1040 (51.1) |  |
| >50% | 933 (45.8) |  |
| Left Ventricular Internal Diameter in Diastole (LVIDd), cm, mean (SD) | 5.48 (0.98) | 3.0 |
| Right Ventricular Function, n (%) |  | 4.3 |
| Normal | 869 (42.7) |  |
| < Severe | 812 (39.9) |  |
| Severe | 268 (13.2) |  |
| RAP, n (%) |  |  |
| Normal [<5 mmHg] | 712 (35.0) |  |
| < Severe [5-15 mmHg] | 594 (29.2) |  |
| Severe [>15 mmHg] | 549 (27.0) |  |
| Missing | 181 (8.9) |  |
| RVSP, mmHg, median (IQR) | 43 (35.0–53.0) | 25.8 |

ACEI, angiotensin converting enzyme inhibitor; ALT, alanine aminotransferase; ARB, angiotensin II receptor blocker; ARNI, angiotensin receptor-neprilysin inhibitor; BMI, body mass index; Bpm, beats per minute; COPD, chronic obstructive pulmonary disease; CRT, cardiac resynchronization therapy; GGT, gamma-glutamyl transferase; ICD, implantable cardioverter-defibrillator; IQR, interquartile range; LVEF, left ventricular ejection fraction; MRA, mineralocorticoid receptor antagonist; NT-proBNP, N-terminal-pro B-type natriuretic peptide; RAP, right atrial pressure; RVSP, right ventricular systolic pressure; SD, standard deviation; SGLT2, sodium-glucose cotransporter 2 inhibitor; WBC, white blood cell.

Table 2 presents the characteristics of the final variables selected by LASSO for the development and validation cohorts. Demographic and clinical characteristics were similar between cohorts. The proportion of 6-month mortality was 29% and 31.1% in the development and validation cohorts, respectively.

**Table 2.** Comparison of Development and Validation Cohorts: Final Variables Used to Calculate the CARDIO-PAL Score.

| **Variable** | **Development**<br>**Cohort (N = 2036)** | **Validation**<br>**Cohort (N = 508)** |
| --- | --- | --- |
| Age (years), mean (SD) | 74 (14.4) | 74 (14.7) |
| Admission PPSv2 Score, mean (SD) | 59 (14.7) | 59 (14.5) |
| Triage SBP (mmHg), mean (SD) | 131.9 (28.4) | 129 (26.8) |
| ACEI/ARB, n (%) |  |  |
| Never | 641 (31.5) | 161 (31.7) |
| Discontinued | 363 (17.8) | 104 (20.5) |
| Current use | 1,031 (50.6) | 243 (47.8) |
| Beta-Blocker, n (%) |  |  |
| Never | 385 (18.9) | 95 (18.7) |
| Discontinued | 229 (11.3) | 58 (11.4) |
| Current use | 1,421 (69.8) | 355 (69.9) |
| MRA, n (%) |  |  |
| Never | 1,402 (68.9) | 365 (71.9) |
| Discontinued | 94 (4.6) | 22 (4.3) |
| Current use | 539 (26.5) | 121 (23.8) |
| Urea, mmol/L, median (IQR) | 10 (6.7–16.6) | 11 (6.7–16.7) |
| RVSP, median (IQR) | 43 (35.0–53.0) | 44 (37.0–55.0) |
| 6-month mortality, n (%) | 590 (29.0) | 158 (31.1) |

ACEI, angiotensin converting enzyme inhibitor; ARB, angiotensin II receptor blocker; IQR, interquartile range; MRA, mineralocorticoid receptor antagonist; PPSv2, Palliative Performance Scale version 2; RVSP, right ventricular systolic pressure; SBP, systolic blood pressure; SD, standard deviation.

### Model Derivation

Eight out of 53 candidate variables were retained through LASSO regression as the most predictive of 6-month mortality. Table 3 represents results from the multivariable logistic regression analysis of the final 8 variables. Key predictors with over twice the odds of predicting 6-month mortality included admission PPsv2 per 10% decrease from 80% (aOR 2.08, 95%CI 1.88-2.23), discontinued use of ACEi/ARB (aOR 2.58 95%CI 1.86-3.57), discontinued use of MRA (aOR 2.67, 95%CI 1.54-4.62), logarithm of RVSP (aOR 2.06, 95%CI 1.41-3.01) and logarithm of urea (aOR 2.13, 95%CI 1.73-2.61). Triage SBP per 10 mmHg decrease from 130 mmHg (aOR 1.14, 95%CI 1.09-1.19) and discontinued use of beta-blocker (aOR 1.59, 95%CI 1.04-2.42) were other important variables included in the final CARDIO-PAL score.

**Table 3.** Multivariable Logistic Regression Models for Predicting 6-month Mortality in the Development Cohort.

| Variable | Full Model $\beta$ | OR (95% CI) | Reduced Model<br>(Excluding RVSP) $\beta$ | OR (95% CI) |
| --- | --- | --- | --- | --- |
| Age (years) | 0.0090 | 1.01 (1.00–1.02) | 0.0085 | 1.01 (1.00–1.02) |
| Admission PPSv2 (per 10% decrease) | 0.7310 | 2.08 (1.89 – 2.33) | 0.7500 | 2.13 (1.85-2.33) |
| Triage SBP (per 10 mmHg decrease) | 0.1315 | 1.14 (1.09-1.19) | 0.1359 | 1.15 (1.10-1.20) |
| ACEI/ARB |  |  |  |  |
| Never (reference) |  |  |  |  |
| Discontinued | 0.6209 | 2.58 (1.86–3.57) | 0.6254 | 2.62 (1.89–3.62) |
| Current use | -0.2941 | 1.03 (0.78–1.37) | -0.2884 | 1.05 (0.79–1.39) |
| Beta-Blocker |  |  |  |  |
| Never (reference) |  |  |  |  |
| Discontinued | 0.3501 | 1.59 (1.04–2.42) | 0.3545 | 1.56 (1.03–2.38) |
| Current use | -0.2394 | 0.88 (0.65–1.19) | -0.2632 | 0.84 (0.62–1.14) |
| MRA |  |  |  |  |
| Never (reference) |  |  |  |  |
| Discontinued | 0.6574 | 2.67 (1.54–4.62) | 0.6815 | 2.76 (1.59–4.79) |
| Current use | -0.3331 | 0.99 (0.74–1.33) | -0.3461 | 0.99 (0.74–1.33) |
| log(RVSP, mmHg) | 0.7235 | 2.06 (1.41–3.01) |  |  |
| log(Urea, mmol/L) | 0.7545 | 2.13 (1.73–2.61) | 0.7997 | 2.23 (1.81–2.73) |

ACEI, angiotensin converting enzyme inhibitor; ARB, angiotensin II receptor blocker; CI, confidence interval; MRA, mineralocorticoid receptor antagonist; OR, odds ratio; PPSv2, Palliative Performance Scale version 2; RVSP, right ventricular systolic pressure; SBP, systolic blood pressure.

Table 4 presents the point assignment and final scoring system used to derive the CARDIO-PAL score. Assigned point values ranged from −1 to +3, with higher scores corresponding to a higher predicted risk of 6-month mortality. Predictors with a 3-point assignment and higher risk of short-term mortality included PPsv2, discontinued use of MRA, log-transformed urea and log-transformed RVSP. Predictors associated with lower risk of short-term mortality and assigned to −1 point included current use of ACEi/ARB, beta-blocker, or MRA, and SBP above 130 mmHg.

**Table 4.**
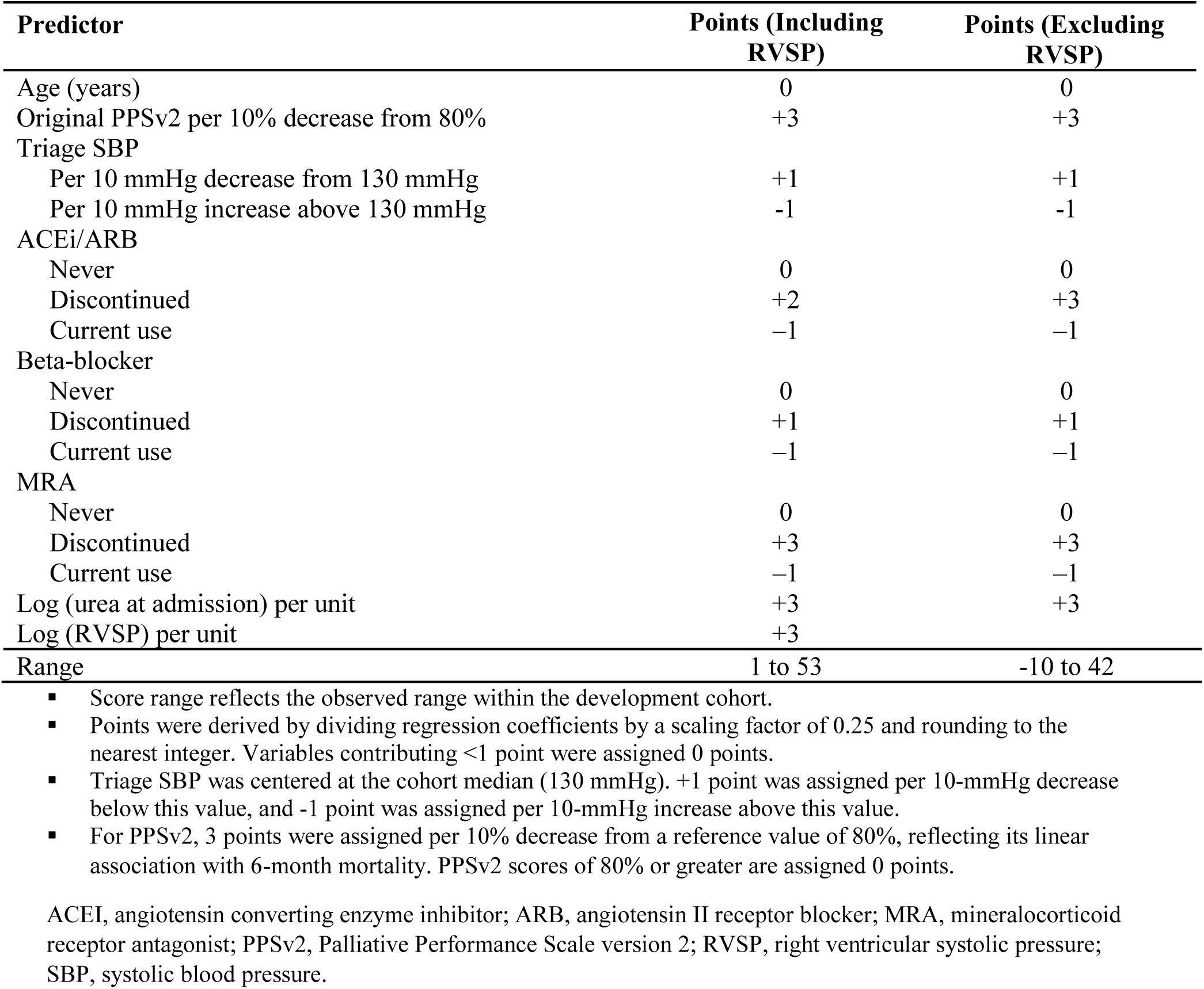
Derivation of the CARDIO-PAL Point Score.

| Predictor | Points (Including RVSP) | Points (Excluding RVSP) |
| --- | --- | --- |
| Age (years) | 0 | 0 |
| Original PPSv2 per 10% decrease from 80% | +3 | +3 |
| Triage SBP |  |  |
| Per 10 mmHg decrease from 130 mmHg | +1 | +1 |
| Per 10 mmHg increase above 130 mmHg | -1 | -1 |
| ACEi/ARB |  |  |
| Never | 0 | 0 |
| Discontinued | +2 | +3 |
| Current use | -1 | -1 |
| Beta-blocker |  |  |
| Never | 0 | 0 |
| Discontinued | +1 | +1 |
| Current use | -1 | -1 |
| MRA |  |  |
| Never | 0 | 0 |
| Discontinued | +3 | +3 |
| Current use | -1 | -1 |
| Log (urea at admission) per unit | +3 | +3 |
| Log (RVSP) per unit | +3 |  |
| Range | 1 to 53 | -10 to 42 |

- Score range reflects the observed range within the development cohort. - Points were derived by dividing regression coefficients by a scaling factor of 0.25 and rounding to the nearest integer. Variables contributing <1 point were assigned 0 points. - Triage SBP was centered at the cohort median (130 mmHg). +1 point was assigned per 10-mmHg decrease below this value, and -1 point was assigned per 10-mmHg increase above this value. - For PPSv2, 3 points were assigned per 10% decrease from a reference value of 80%, reflecting its linear association with 6-month mortality. PPSv2 scores of 80% or greater are assigned 0 points.
ACEI, angiotensin converting enzyme inhibitor; ARB, angiotensin II receptor blocker; MRA, mineralocorticoid receptor antagonist; PPSv2, Palliative Performance Scale version 2; RVSP, right ventricular systolic pressure; SBP, systolic blood pressure.

The final CARDIO-PAL score ranged from 1 to 53 when RVSP was included and −10 to 42 when RVSP was excluded.

Observed 6-month mortality across CARDIO-PAL risk groups was 2%, 15%, and 54% for CARDIO-PAL scores of ≤ 17 (low risk), 18-24 (intermediate risk), and ≥ 25 (high risk), respectively (Table 5; Figure 2). When RVSP was excluded from the model, observed 6-month mortality across risk groups was 2%, 12%, and 52% for CARDIO-PAL score ranges of ≤ 5 (low risk), 6–12 (intermediate risk), and ≥ 13 (high risk), respectively. Figure 3 shows cumulative mortality following hospital discharge over 4 years by CARDIO-PAL risk group. At 4 years, cumulative mortality was 28.6%, 55.6%, and 83.5% in the low-, intermediate-, and high-risk groups, respectively, with clear separation between risk groups evident within the first 180 days.

**Figure 2:**
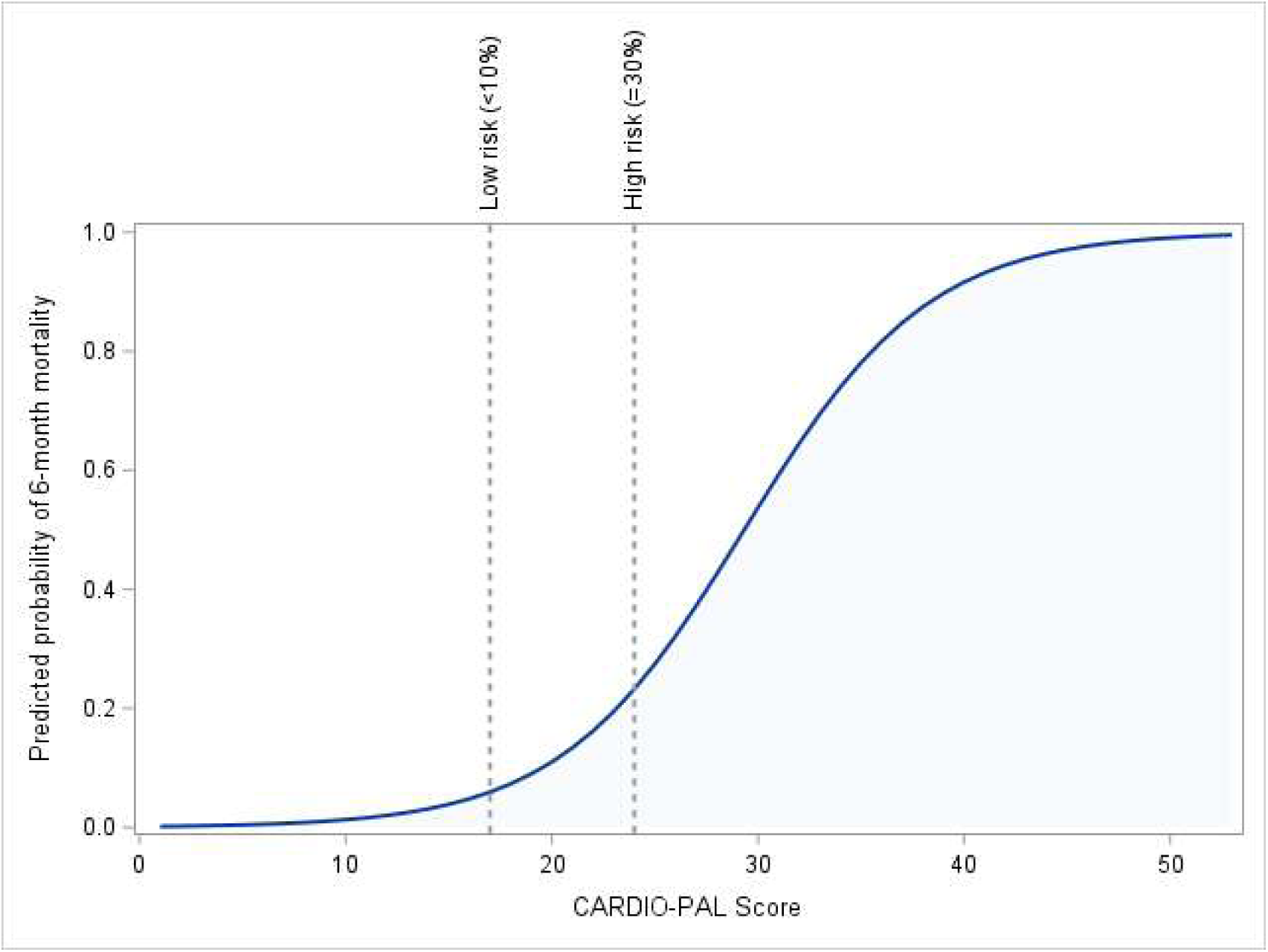
Predicted probability of 6-month mortality by CARDIO-PAL score (development cohort). Vertical dashed lines indicate thresholds for low (<10%) and high (≥30%) risk, with intermediate risk (10–30%) between these values.

**Figure 3.**
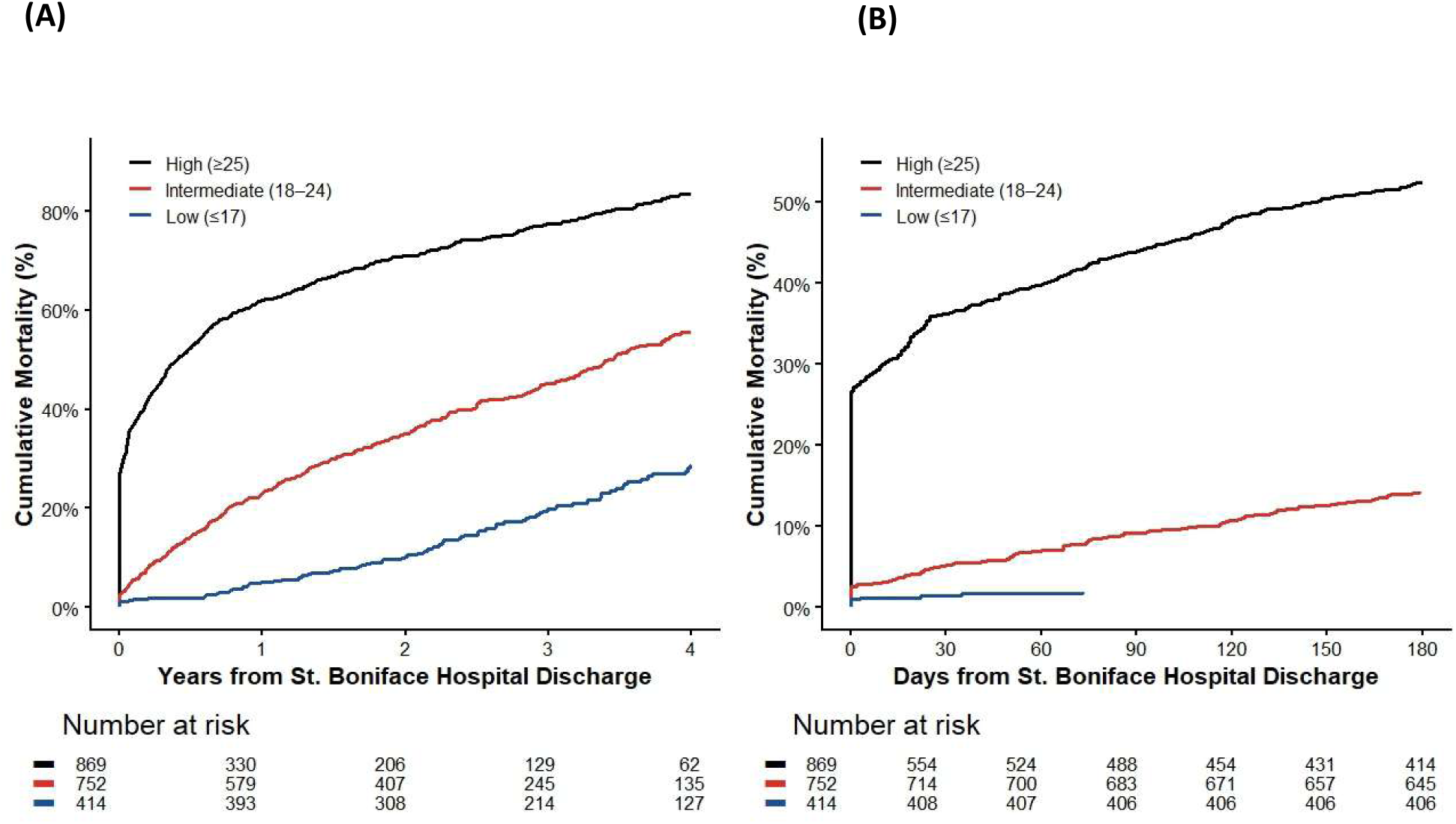
Kaplan–Meier estimates of all-cause mortality following discharge from the tertiary care hospital (St. Boniface Hospital) according to CARDIO-PAL risk group. (A) Cumulative mortality over 4 years following discharge. (B) Magnified view of the first 180 days of follow-up using the same Kaplan– Meier estimates shown in panel (A). Follow-up was administratively censored on April 7, 2025, for patients who remained alive. Patients were classified as low (≤17 points), intermediate (18–24 points), or high (≥25 points) risk based on their CARDIO-PAL score. Numbers at risk are shown below the x-axis.

**Table 5.** Risk Classification Groups for the CARDIO-PAL Score.

| **Risk Group** | **With RVSP**<br>**Score** | **Mortality**<br>**(%)** | **Without RVSP**<br>**Score** | **Mortality**<br>**(%)** |
| --- | --- | --- | --- | --- |
| I (Low) | ≤17 | 2 | ≤5 | 2 |
| II (Intermediate) | 18–24 | 15 | 6–12 | 12 |
| III (High) | ≥25 | 54 | ≥13 | 52 |

Observed 6-month mortality after grouping patients by their CARDIO-PAL score. Risk groups were defined based on predicted probability ranges (<10%, 10–30%, ≥30%) and corresponding CARDIO-PAL score ranges derived from the development cohort.
RVSP, right ventricular systolic pressure.

### Model Performance

The CARDIO-PAL score demonstrated strong discrimination in the development cohort (AUC 0.839, 95% CI 0.82-0.86) with minimal overfitting after bootstrap correction (optimism-corrected AUC 0.837, calibration slope 0.98, Table 6). Performance remained stable in the independent validation cohort (AUC 0.80, 95% CI 0.75-0.84), with acceptable calibration (slope 0.84, intercept -0.04, Figure 4).

**Figure 4:**
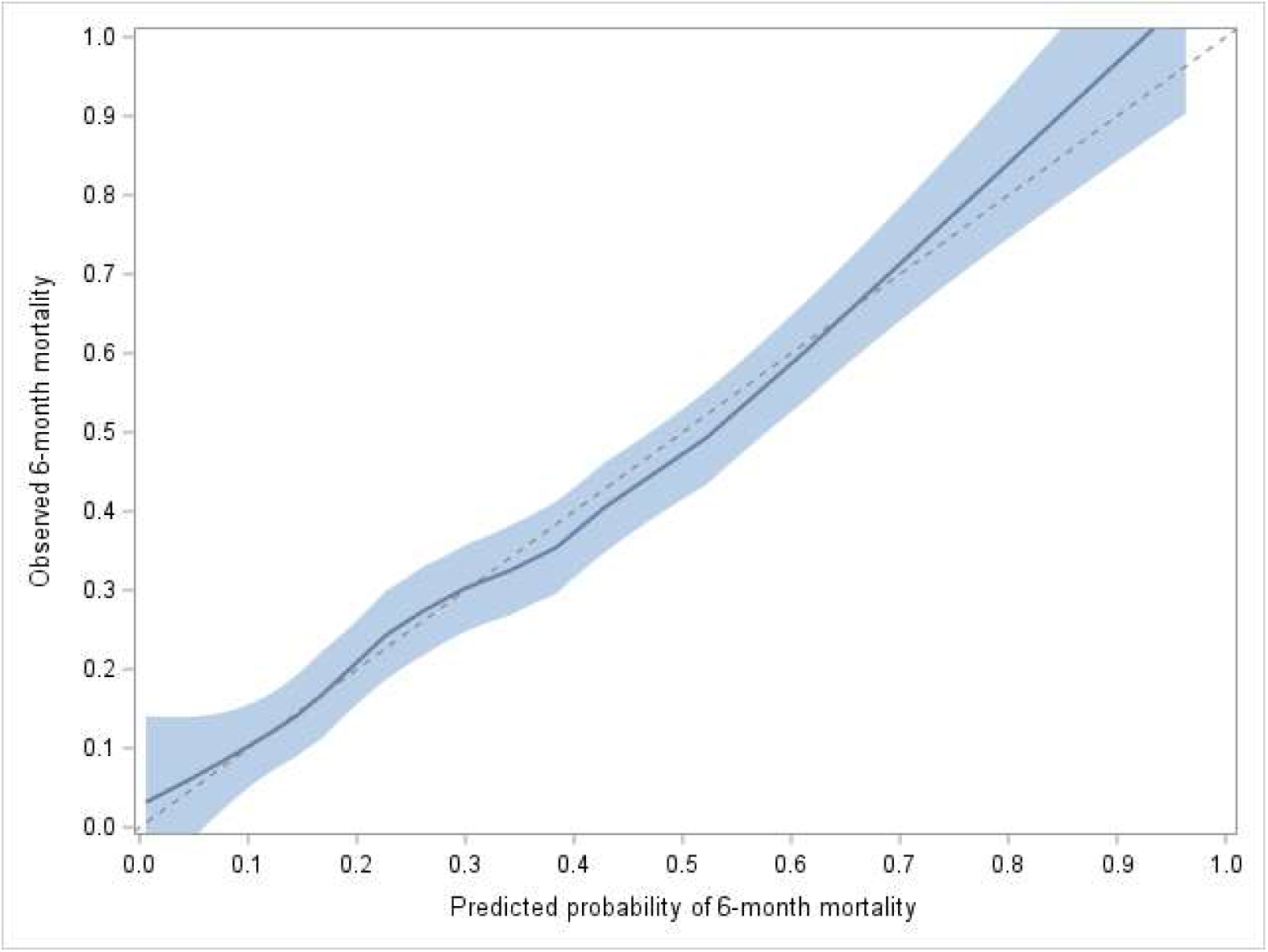
Calibration plot for the CARDIO-PAL score in the validation cohort (N=508). The LOESS-smoothed curve demonstrates the relationship between predicted and observed 6-month mortality. The shaded area represents the 95% confidence interval, and the dashed line indicates perfect calibration.

**Table 6.** Discrimination and Calibration of the CARDIO-PAL Score in the Development and Validation Cohorts.

| **Performance Measure** | **Apparent Development**<br>**(80%) N=2036** | **Optimism-Corrected**<br>**(Bootstrap)** | **Holdout (20%)**<br>**[validation]**<br>**N=508** |
| --- | --- | --- | --- |
| AUC (95% CI) | 0.839 (0.82–0.86) | 0.837 | 0.797 (0.75-0.84) |
| Calibration Slope | 1 | 0.98 | 0.84 |
| Calibration Intercept | 0 | — | –0.04 |

Apparent performance reflects model performance in the development cohort. Optimism-corrected estimates were obtained using 1,000 bootstrap resamples. The optimism-corrected calibration slope represents the shrinkage factor (1 – mean optimism). Validation results are from the independent 20% holdout cohort.
AUC, area under the curve; CI, confidence interval.

Measures of diagnostic accuracy were evaluated in the validation cohort (Table 7). Using a pre-specified predicted probability threshold of ≥0.55 for 6-month mortality to define the high-risk group, sensitivity, specificity, PPV and NPV for the CARDIO-PAL score were 37%, 91%, 64% and 76% respectively. Similar values were observed when RVSP was excluded from the model (Table 7).

**Table 7.** Clinical Threshold Performance of CARDIO-PAL in the Validation Cohort.

| Performance Measure | Original PPSv2 | CARDIO-PAL | CARDIO-PAL (without RVSP) |
| --- | --- | --- | --- |
| Sensitivity | 36.70% | 43.00% | 43.67% |
| Specificity | 90.60% | 93.10% | 93.71% |
| Positive Predictive Value | 63.70% | 73.90% | 75.82% |
| Negative Predictive Value | 76.00% | 78.40% | 78.66% |

High-risk classification for the original PPSv2 was defined as a score $\leq 40$ . For CARDIO-PAL, high risk was defined as a predicted probability $\geq 0.55$ , selected a priori in the derivation cohort to prioritize high specificity.
PPSv2, Palliative Performance Scale version 2; RVSP, right ventricular systolic pressure.

## Discussion

The CARDIO-PAL score is a novel prognostic tool, developed and validated to predict 6-month mortality in HF inpatients. The score is practical, relying on 8 easily attainable variables including age, PPSv2, blood pressure, medication use, urea, and RVSP. A version of CARDIO-PAL excluding RVSP was also created, to acknowledge the fact that RVSP is not always quantifiable and to avoid the need to obtain an echocardiogram strictly for the purpose of prognostication. Performance of the model was strong with good discrimination (AUC = 0.80) in the validation cohort and predicted mortality risk closely matching observed outcomes.

Well known prognostic models, such as the Meta-Analysis Global Group in Chronic Heart Failure (MAGGIC) score and Seattle Heart Failure Model (SHFM) predict death at one year and beyond.^14,15^ This limits usefulness when discussing referral to palliative care and hospice programs, many of which use <6-month prognosis as enrollment criteria.^16^ Allen et al. estimated the probability of death in 10930 ambulatory patients with HF using SHFM and MAGGIC.^17^ SHFM predicted a >50% chance of 1-year mortality in only 8/1661 patients who died (sensitivity 0.5%),^17^ while 60% of patients who actually died were only predicted to have a <15% chance of death.^17^ Similarly, MAGGIC predicted a >50% chance of 1-year mortality in only 52/1661 of the patients that died (sensitivity 3.1%) while 55% of patients who actually died were only predicted to have a <25% chance of death.^17^ Both models substantially underestimated mortality among patients who subsequently died, highlighting a lack of predictability at the individual patient level and underscoring the need for an accurate, individualized short-term mortality risk score.

Variables used to represent health fall on a continuum of objective biologic variables, such as laboratory or biomarker values, to more complex and subjective measures such as physical function and health perception.^18^ Most available HF risk scores combine objective variables to estimate mortality. While objective, these laboratory-based predictive scores fail to capture more nuanced and personalised information and do not consider a patient in their entirety, possibly explaining the breakdown in predictive accuracy at an individual level.^17,18^ One risk score which does include subjective variables is the EPICTER score, which was developed to predict 6-month mortality in HF inpatients.^19^ It incorporates estimated ≤6-month mortality and patient or family acceptance of a palliative approach.^19^ Although perceived health is well known to predict mortality, the lack of objective and reproducible methods limits its use.^18^ In contrast, CARDIO- PAL aims to incorporate features across the health spectrum. It integrates biology-based values, such as urea and SBP, with a functional subjective measure (PPSv2), in order to provide a holistic predictive model. While subjective, PPSv2 has the benefit of being a standardized measure of function, imparting reliability and validity.

A major strength of CARDIO-PAL is the inclusion of an assessment of function by way of the PPSv2. The PPSv2 is simple and quick to use, taking less than 2 minutes to assign (Supplementary Figure 1). Component information is readily available from the review of standard nursing records, making retrospective abstraction or abstraction using artificial intelligence possible. The PPSv2 emerged as the strongest predictor of short-term mortality amongst the variables assessed (OR 2.08 per 10% decrease in PPSv2, contributing 3 points to the score per 10% decrease from 80%). The PPSv2 serves as a powerful surrogate for, and strongly overlaps with, assessments of frailty, such as the Fried phenotype and Rockwood’s accumulation of deficits model, both of which are strongly linked to mortality.^20,21^ Despite this, many HF risk scores do not include measures of functional status, with a few exceptions.^6,19,22–24^ Those that do include functional status measures usually only include a single aspect of physical function such as the New York Heart Association (NYHA) class or the 6-minute walk test (6MWT). In contrast, the PPSv2 provides a more comprehensive evaluation of global functional status by incorporating multiple domains, including ambulation, activity level, self-care, oral intake, and level of consciousness.

By design, the variables chosen to study were considered clinically relevant and/or were components of previously published HF prognostic scores. As a result, 36/53 (68%) variables were statistically associated with ≤ 6-month mortality (p<0.05, Supplementary Figure 2). After LASSO regression, the most predictive variables were selected to create a parsimonious model to avoid overfitting and serve as a practical tool for clinicians. The most predictive variables were: PPSv2, SBP (hypotension), cardiac medication use, urea, and RVSP.

1. Hypotension is multifactorial, relating to a low output state, severe valvular disease, hyponatremia, and/or low blood volume.^25^ It is a common variable included in HF risk scores, due to its known association with HF severity and mortality.^25^
2. Medication (ACEi/ARB, beta-blocker, and MRA) discontinuation is not random, and is driven by frailty, hypotension, and multiorgan involvement, all markers of advanced disease.^2,26,27^ Discontinuation of cardiac medications has a higher mortality rate than never being on the medication, suggesting this is not due to medication effects nor a result of neglecting guideline directed medical therapy. Underlying features of poor health prompting appropriate discontinuation are more likely the reason.
3. Urea, a marker of renal dysfunction, and RVSP, a surrogate for pulmonary hypertension, reflect progressive and advanced HF.^27^ A version of CARDIO-PAL which does not include RVSP was created to improve translatability. It acknowledges that RVSP is not always quantifiable and prevents the need to order an echocardiogram solely for prognostic purposes, in cases where it does not align with a patient’s goals of care.
4. Age was integrated into the CARDIO-PAL model as an important demographic variable and is a component of most available HF prognostic scores.^6^ Although age provides zero points in the CARDIO-PAL score, it impacts the relative point distribution among the other variables. In previous scoring systems, the importance of age is to serve as a surrogate for frailty. Therefore, by using a more powerful and comprehensive surrogate in CARDIO-PAL (e.g. functional status such as PPSv2, or inability to tolerate medications), chronological age becomes less important. This concept is reflected in many palliative care tools.^28,29^

Published prognostic models to predict short term (≤ 6-month) mortality in the HF population have been previously reviewed.^6^ The literature is heterogeneous, spanning cardiology, geriatric medicine, and palliative care disciplines, with scores developed in vastly different HF or mixed non-malignant patient cohorts.^6^ Additionally, short term prognostic scores were limited by high risk of bias due to a lack of robust methodology and concern over applicability when applied through the lens of palliative care decision making.^6^ CARDIO-PAL was developed from an inclusive multimorbid HF cohort, to reflect a real-world population. CARDIO-PAL performance (AUC = 0.8) was comparable to the most discriminatory cardiology model, EHMRG, which predicts 1-week mortality in HF patients presenting to the emergency department (AUC = 0.83).^30^ Furthermore, CARDIO-PAL proved more discriminatory than models developed to inform palliative care referral decision making, such as EPICTER (AUC = 0.68), PPI (AUC = 0.69), PALIAR (AUC = 0.69), and MPI (AUC = 0.80).^19,28,31,32^

A gold standard to predict mortality in HF patients with perfect accuracy does not exist and likely will never exist. While an attempt was made to directly compare the performance of CARDIO- PAL with other HF risk scores using our dataset, the majority of scores were not developed to predict 6-month mortality, precluding a direct comparison (Table 8).^22,30,33–38^ Of those that were designed to predict 6-month mortality, comparison was not possible due to the need for NT-proBNP at discharge (too few discharge values in our dataset; ELAN-HF), inability to collect estimates of 6-month mortality or acceptance of palliative care by retrospective review (EPICTER), or use of BNP which was not available at our institution (ESCAPE).^19,23,24^ An increased focus on how to collaborate and integrate multiple tools (i.e. clinical features, diagnostic results, and functional status), alongside clinical judgement, would better inform HF patients and prioritize patient-centered care. CARDIO-PAL, being a more specific (93%) but less sensitive (43%) predictive tool than other direct comparators (Table 9), can add valuable information to this approach by minimizing “false positives” in the high-risk group. As high-risk patients may be considered for intensification of guideline directed treatment, advanced therapies, and/or referral to palliative care in keeping with patient wishes misclassification in this group could have negative consequences on appropriate treatments, patient psychosocial wellbeing, and health care resources. A collaborative approach, integrating multiple prognostic tools is one way to address the unpredictable nature of chronic organ failure. As an example, a low CARDIO-PAL score at one moment does not equate to guaranteed survival, as for every 100 low-risk patients in CARDIO- PAL, 20 (20%) will die at 4-years. Conversely, for every 100 high-risk patients, 20 (20%) will still be alive at 4 years. Prognostic scores often breakdown for multiple reasons, such as: natural difficulty predicting cardiac death (which includes sudden death), death due to competing comorbidities that are not measured in HF prognostic tools (e.g. extent of metastatic cancer, severity of a neurological disorder, state of other organ failures), psychosocial variables that are difficult to quantify (e.g. unstable housing, social isolation, limited caregiver support), or very specific patient circumstances that cannot be well accounted for in prognostic models meant for a generalized population (e.g. rare congenital cardiac disease). For this reason, a holistic model must also include clinical judgement alongside risk estimates. It is important for clinicians to follow patients, examine trajectory changes over time, and account for individualized characteristics of advanced HF. Truthful conversation with patients and families should highlight the concept of hoping for the best, while planning for the worst. Ultimately, the CARDIO-PAL score is a tool not to be used in isolation, but as a component of a global and holistic patient centered assessment, alongside other risk estimates of prognosis and clinical assessments.

**Table 8:**
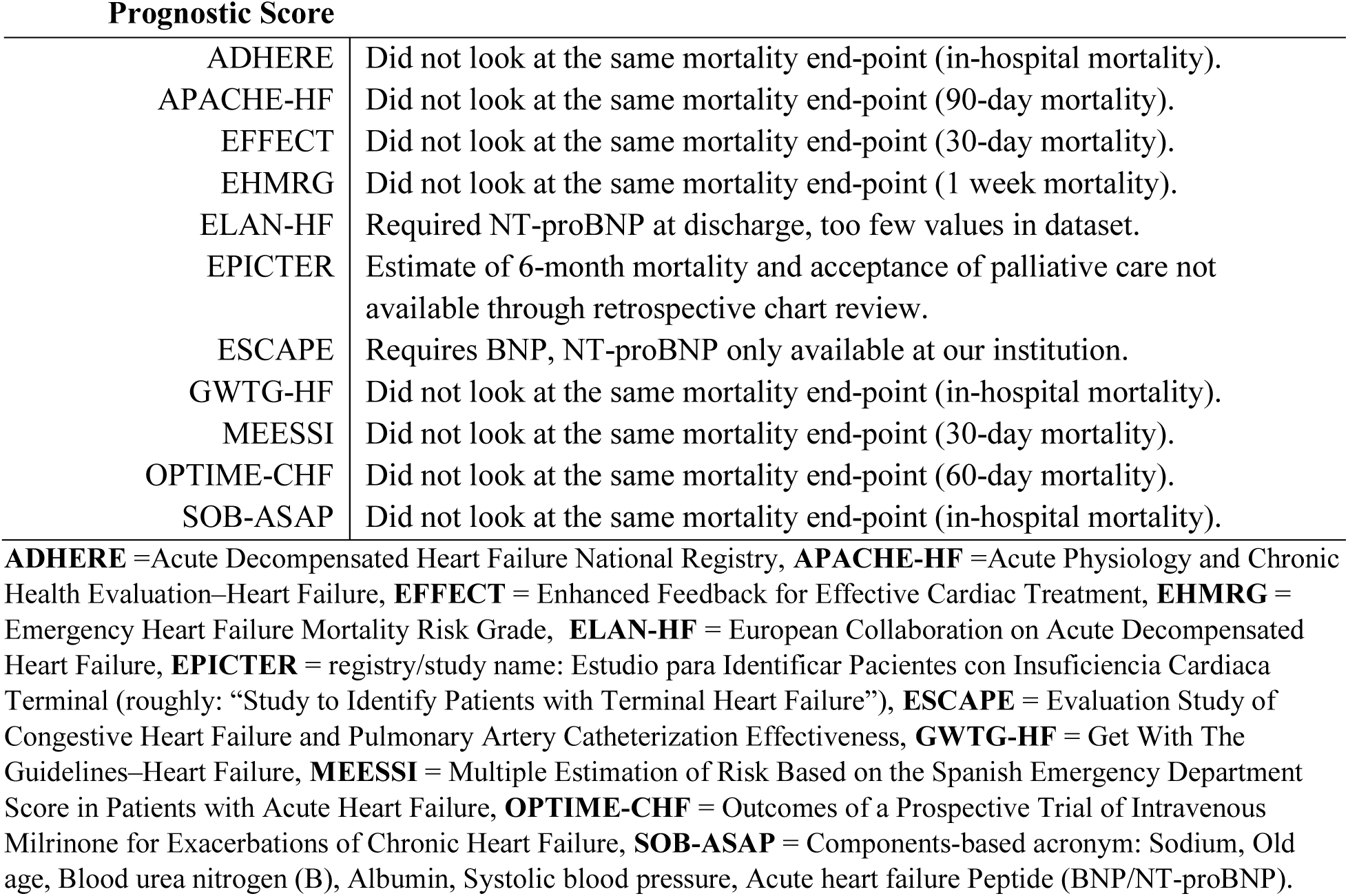
Specifics regarding the inability to compare the performance of CARDIO-PAL with other HF prognostic risk scores developed to predict short term mortality.

**Table 9:** Comparison of test characteristics between CARDIO-PAL and previously derived scoring systems to predict 6-month mortality in heart failure patients.

|  | **SENSITIVITY** | **SPECIFICITY** | **POSITIVE**<br>**PREDICTIVE**<br>**VALUE** | **NEGATIVE**<br>**PREDICTIVE**<br>**VALUE** | **6-MONTH**<br>**MORTALITY** |
| --- | --- | --- | --- | --- | --- |
| **CARDIO-PAL** | **43%** | **93.1%** | **73.9%** | **78.4%** | **31.1%** |
| ELAN-HF | Not reported | Not reported | Not reported | Not reported | 15% |
| EPICTER | 68.8% | 61.8% | 41% | 83.7% | 26.9% |
| ESCAPE | Not reported | Not reported | Not reported | Not reported | 18.7% |

**APACHE-HF** =Acute Physiology and Chronic Health Evaluation–Heart Failure, **ELAN-HF** = European Collaboration on Acute Decompensated Heart Failure, **EPICTER** = registry/study name: Estudio para Identificar Pacientes con Insuficiencia Cardiaca Terminal (roughly: “Study to Identify Patients with Terminal Heart Failure”), **ESCAPE** = Evaluation Study of Congestive Heart Failure and Pulmonary Artery Catheterization Effectiveness.

This study was limited by factors common to all retrospective designs, and the accuracy of information was dependent on documentation at the time of the patient encounter. Secondly, there was high missingness of certain variables. This reflects the real-world population chosen and the practice of de-escalation as patients near the end of life. Thirdly, PPSv2 was only collected closest to admission. Although, this is standard in palliative care scoring tools,^7^ it does not capture whether a change in PPSv2 over the course of admission predicts mortality. While this would have been interesting, longitudinal changes in predictive variables are far beyond the scope of this study. Notably, other prognostic models did not incorporate changes in predictor variables over time. However, the overwhelming predictability of admission PPSv2 itself is an important finding. Finally, validation in a prospective and external cohort, as well as outpatient population, would further improve the generalizability of the score.

## Conclusions

The CARDIO-PAL score is a novel and practical tool to predict 6-month mortality in HF inpatients, specifically developed to help identify patients for palliative care or hospice referral. It includes an easy-to-use comprehensive functional assessment (PPSv2), which is highly predictive of mortality. We challenge care providers to integrate CARDIO-PAL with the information provided by other prognostic tools and clinical judgement, in order to best apply a collaborative and holistic approach to patients with advanced HF. By identifying HF patients at risk of short-term death, an integrative approach including CARDIO-PAL, can help to prioritize patients for timely goals of care conversations and facilitate referral to palliative care and hospice programs.

## ABBREVIATIONS AND ACRONYMS

CARDIO-PAL =: Cardiac Assessment of Risk to Determine Illness Outcomes and Palliative Care
EPICTER =: estimates of 6-month mortality or acceptance of palliative care by retrospective review
HF =: heart failure
LVAD =: left ventricular assist device
MAGGIC =: Meta-Analysis Global Group in Chronic Heart Failure
MAID =: medical assistance in dying
NT-proBNP =: N-terminal pro-B-type natriuretic peptide
PPSv2 =: Palliative Performance Scale version 2
SHFM =: Seattle Heart Failure Model

## Data Availability

Data referred to in the manuscript is available upon request.

---

**Supplementary Figure S1:**
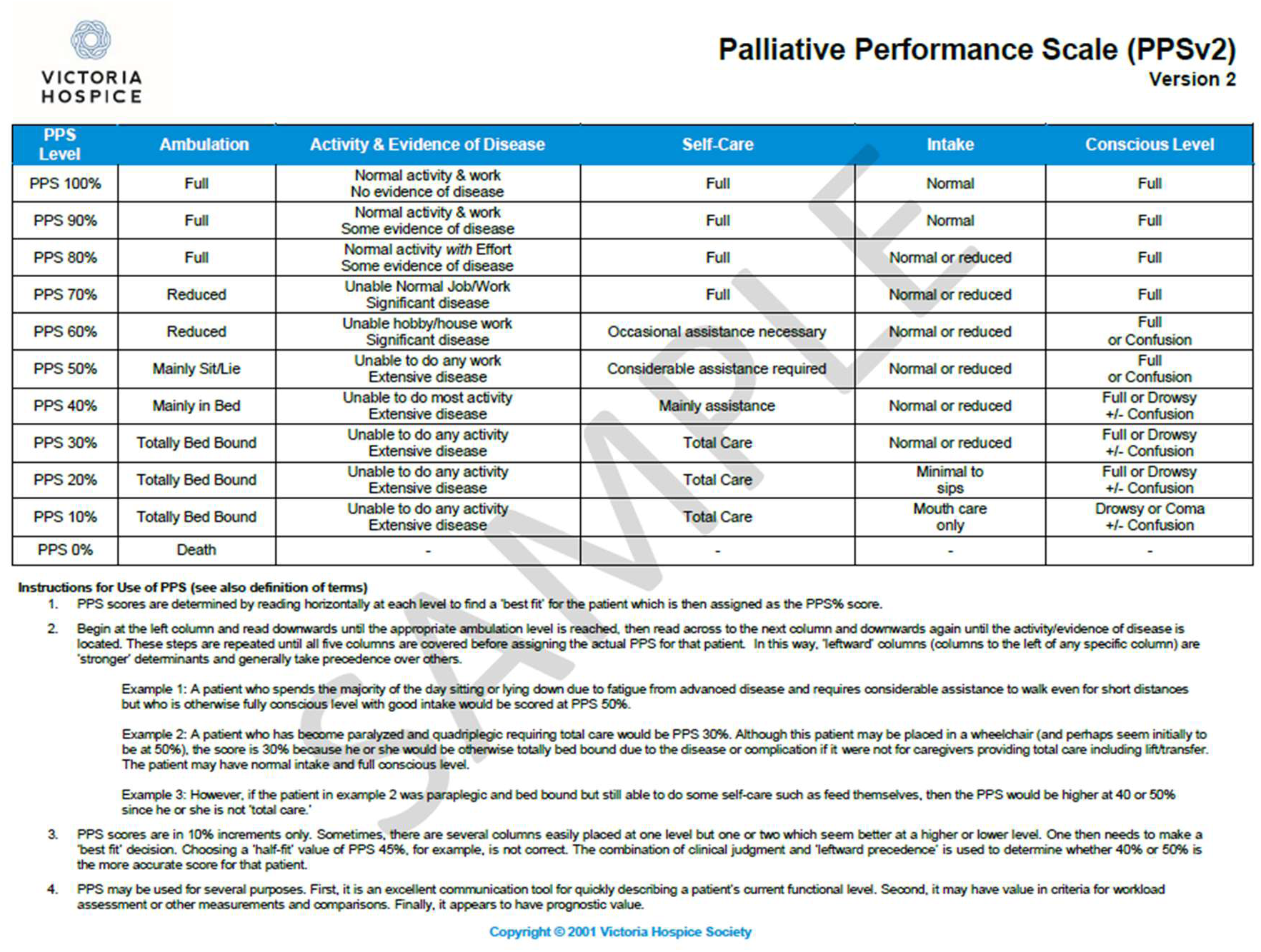
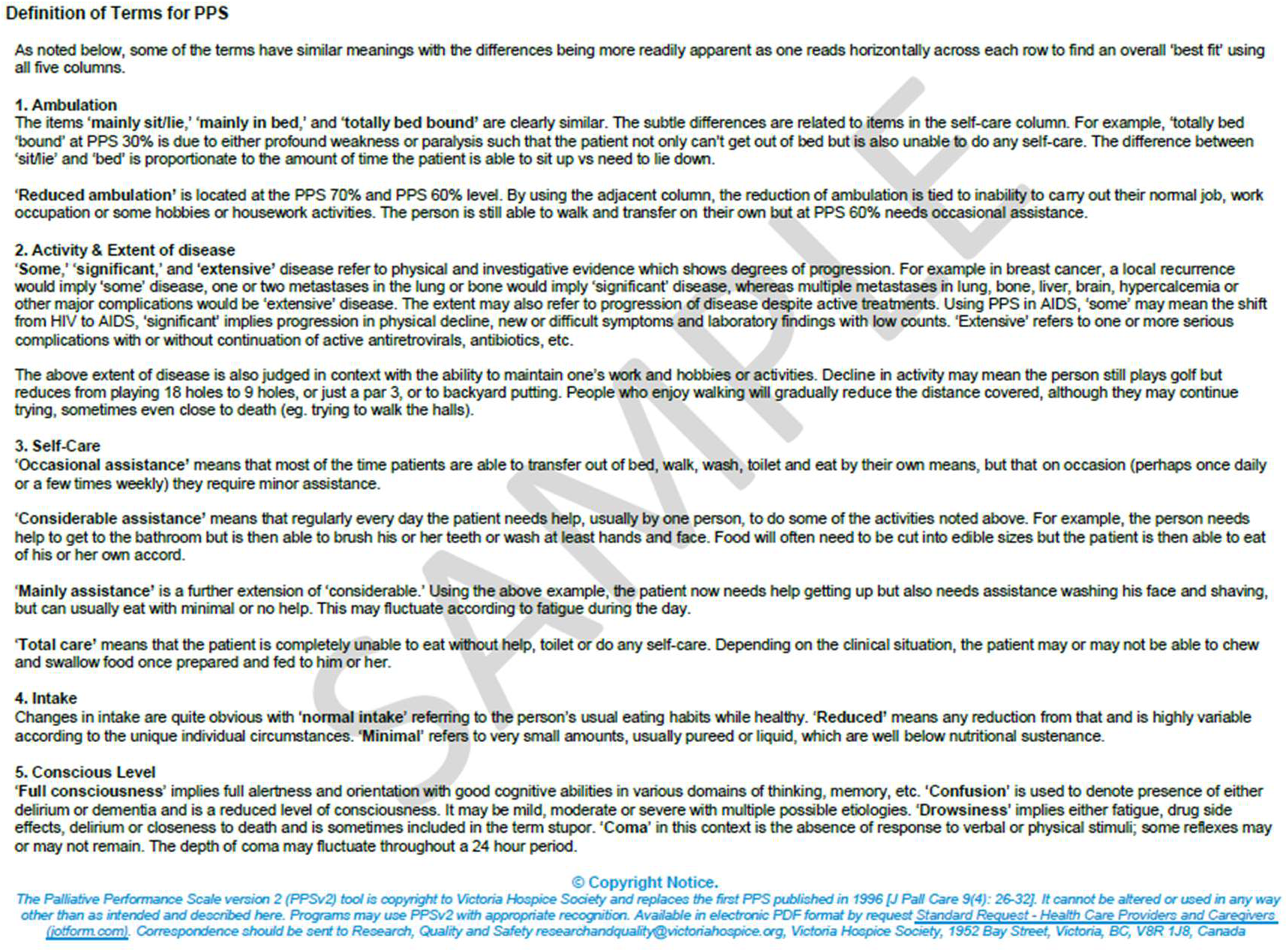
The Palliative Performance Scale (PPSv2). Written permission to use the PPSv2 was obtained from Victoria Hospice.

**Supplementary Figure S2.**
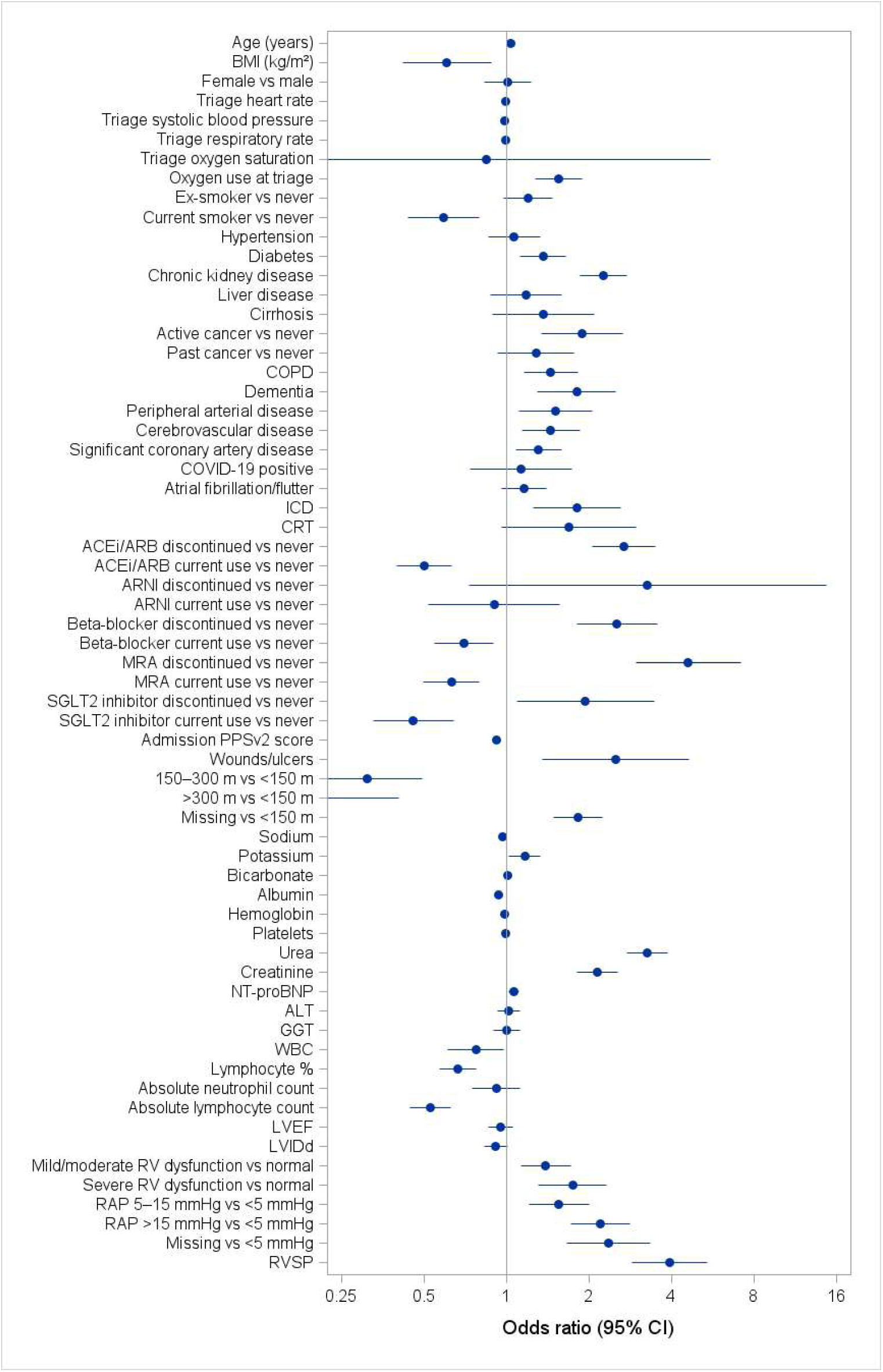
Forest plot of univariable associations between candidate predictors and 6-month mortality For binary variables, odds ratios represent the presence vs absence of the characteristic. 36/53 (68.0%) candidate variables were found statistically significant and associated with 6-month mortality at p<0.05. LASSO regression was subsequently used to select the most parsimonious model and minimize overfitting.

## Acknowledgments

The authors would like to thank Yuping Cao for her help with manuscript preparation.

## References

1. Walsh MN, Kober L, Sliwa K, et al. AHA/ACC/ESC/WHF Expert Consensus Document: Second Universal Definition of Heart Failure (2026). Circulation. 2026;154:e279–e293..

2. Steinberg L, White M, Arvanitis J, Husain A, Mak S. Approach to advanced heart failure at the end of life. Can Fam Physician. 2017;63:674–680.

3. Sobanski PZ, Alt-Epping B, Currow DC, et al. Palliative care for people living with heart failure: European Association for Palliative Care Task Force expert position statement. Cardiovasc Res. 2020;116:12–27.

4. Heidenreich PA, Bozkurt B, Aguilar D, et al. 2022 AHA/ACC/HFSA Guideline for the Management of Heart Failure: A Report of the American College of Cardiology/American Heart Association Joint Committee on Clinical Practice Guidelines. Circulation. 2022;145: e895– e1032.

5. McDonagh TA, Metra M, Adamo M, et al. 2021 ESC Guidelines for the diagnosis and treatment of acute and chronic heart failure. Eur Heart J. 2021;42:3599–3726.

6. Whalen K, Bews H, Tam JW, Quan S, Pilkey J. Prognostic Models to Trigger Palliative Care Referrals in Heart Failure: A Scoping Review. CJC Open. 2026. 10.1016/j.cjco.2026.06.011.

7. Masterson Creber R, Russell D, Dooley F, et al. Use of the Palliative Performance Scale to estimate survival among home hospice patients with heart failure. ESC Heart Fail. 2019;6:371–378.

8. Baik D, Russell D, Jordan L, Dooley F, Bowles KH, Masterson Creber RM. Using the Palliative Performance Scale to Estimate Survival for Patients at the End of Life: A Systematic Review of the Literature. J Palliat Med. 2018;21:1651–1661.

9. Sumi T, Umemoto N, Kajiura H, et al. Prognostic utility of Palliative Prognostic Index for prediction of 30-day and 1-year outcome in patients with acute decompensated heart failure. Eur Heart J. 2019;40: ehz745.0942.

10. Nieto Martín MD, Bernabeu Wittel M, de la Higuera Vila L, et al. Adaptation of the Palliative Prognostic Index in patients with advanced medical conditions. Rev Clin Esp. 2013;213:323–329.

11. Collins GS, Reitsma JB, Altman DG, Moons KGM. Transparent reporting of a multivariable prediction model for individual prognosis or diagnosis (TRIPOD): the TRIPOD statement. BMJ. 2015;350:g7594.

12. Cai D, Chen Q, Mu X, et al. Development and validation of a novel combinatorial nomogram model to predict in-hospital deaths in heart failure patients. BMC Cardiovasc Disord. 2024;24:16.

13. Jansen WJJ, Buma S, Gootjes JRG, Zuurmond WWA, Perez RSGM, Loer SA. The palliative performance scale applied in high-care residential hospice: a retrospective study. J Palliat Med. 2015;18:67–70.

14. Pocock SJ, Ariti CA, McMurray JJV, et al. Predicting survival in heart failure: a risk score based on 39 372 patients from 30 studies. Eur Heart J. 2013;34:1404–1413.

15. Levy WC, Mozaffarian D, Linker DT, et al. The Seattle Heart Failure Model: prediction of survival in heart failure. Circulation. 2006;113:1424–1433.

16. Fine PG. Hospice Underutilization in the U.S.: The Misalignment of Regulatory Policy and Clinical Reality. J Pain Symptom Manage. 2018;56:808–815.

17. Allen LA, Matlock DD, Shetterly SM, et al. Use of Risk Models to Predict Death in the Next Year Among Individual Ambulatory Patients With Heart Failure. JAMA Cardiol. 2017;2:435– 441.

18. Cleary PD. Subjective and objective measures of health: which is better when? J Health Serv Res Policy. 1997;2:3–4.

19. Romero-Correa M, Salamanca-Bautista P, Bilbao-González A, et al. The EPICTER Score: A Bedside and Easy Tool to Predict Mortality at 6 Months in Acute Heart Failure. ESC Heart Fail. 2022;9:2348–2356.

20. Matsue Y, Kamiya K, Saito H, et al. Prevalence and prognostic impact of the coexistence of multiple frailty domains in elderly patients with heart failure: the FRAGILE-HF cohort study. Eur J Heart Fail. 2020;22:2112–2119.

21. Rockwood K, Song X, MacKnight C, et al. A global clinical measure of fitness and frailty in elderly people. CMAJ. 2005;173:489–495.

22. Wussler D, Kozhuharov N, Sabti Z, et al. External Validation of the MEESSI Acute Heart Failure Risk Score: A Cohort Study. Ann Intern Med. 2019;170:248–256.

23. O’Connor CM, Hasselblad V, Mehta RH, et al. Triage after hospitalization with advanced heart failure: the ESCAPE (Evaluation Study of Congestive Heart Failure and Pulmonary Artery Catheterization Effectiveness) risk model and discharge score. J Am Coll Cardiol. 2010;55:872– 878.

24. Salah K, Kok WE, Eurlings LW, et al. A novel discharge risk model for patients hospitalised for acute decompensated heart failure incorporating N-terminal pro-B-type natriuretic peptide levels: a European coLlaboration on Acute decompeNsated Heart Failure: ELAN-HF Score. Heart. 2014;100:115–125.

25. Skouri H, Girerd N, Monzo L, et al. Clinical Management and Therapeutic Optimization of Patients with Heart Failure with Reduced Ejection Fraction and Low Blood Pressure. A Clinical Consensus Statement of the Heart Failure Association (HFA) of the ESC. Eur J Heart Fail. 2025;27:707–722.

26. Crespo-Leiro MG, Barge-Caballero E. Advanced Heart Failure: Definition, Epidemiology, and Clinical Course. Heart Fail Clin. 2021;17:533–545.

27. Bozkurt B, Coats AJ, Tsutsui H, et al. Universal Definition and Classification of Heart Failure: A Report of the Heart Failure Society of America, Heart Failure Association of the European Society of Cardiology, Japanese Heart Failure Society and Writing Committee of the Universal Definition of Heart Failure. J Card Fail. 2021; 27:387–413.

28. Nieto Martín MD, Bernabeu Wittel M, de la Higuera Vila L, et al. Adaptation of the Palliative Prognostic Index in patients with advanced medical conditions. Rev Clin Esp. 2013;213:323–329.

29. Hamatani Y, Iguchi M, Ikeyama Y, et al. Comprehensive symptom assessment using Integrated Palliative care Outcome Scale in hospitalized heart failure patients. ESC Heart Fail. 2022;9:1963–1975.

30. Lee DS, Stitt A, Austin PC, et al. Prediction of heart failure mortality in emergent care: a cohort study. Ann Intern Med. 2012;156:767–775, W-261, W-262.

31. Bernabeu-Wittel M, Murcia-Zaragoza J, Hernández-Quiles C, et al. Development of a six-month prognostic index in patients with advanced chronic medical conditions: the PALIAR score. J Pain Symptom Manage. 2014;47:551–565.

32. Huynh BC, Rovner A, Rich MW. Identification of older patients with heart failure who may be candidates for hospice care: development of a simple four-item risk score. J Am Geriatr Soc. 2008;56:1111–1115.

33. Fonarow GC, Adams KF, Abraham WT, Yancy CW, Boscardin WJ, ADHERE Scientific Advisory Committee SG and I. Risk stratification for in-hospital mortality in acutely decompensated heart failure: classification and regression tree analysis. JAMA. 2005;293:572– 580.

34. Lee DS, Austin PC, Rouleau JL, Liu PP, Naimark D, Tu JV. Predicting mortality among patients hospitalized for heart failure: derivation and validation of a clinical model. JAMA. 2003;290:2581–2587.

35. Peterson PN, Rumsfeld JS, Liang L, et al. A validated risk score for in-hospital mortality in patients with heart failure from the American Heart Association get with the guidelines program. Circ Cardiovasc Qual Outcomes. 2010;3:25–32.

36. Shiraishi Y, Kohsaka S, Abe T, et al. Derivation and Validation of Clinical Prediction Models for Rapid Risk Stratification for Time-Sensitive Management for Acute Heart Failure. J Clin Med. 2020;9:3394.

37. Felker GM, Leimberger JD, Califf RM, et al. Risk stratification after hospitalization for decompensated heart failure. J Card Fail. 2004;10:460–466.

38. Okazaki H, Shirakabe A, Hata N, et al. New scoring system (APACHE-HF) for predicting adverse outcomes in patients with acute heart failure: evaluation of the APACHE II and Modified APACHE II scoring systems. J Cardiol. 2014;64:441–449.

